# Statistical Parametric Mapping of Gaze Duration: A Novel Application of a Spatially Extended Statistical Approach to Eye Movement Data

**DOI:** 10.64898/2026.04.30.26351939

**Authors:** Neena Singh, Peter Zeidman, Guillaume Flandin, Pedro Quijada Leyton, Catherine Doogan, Thomas Nyffeler, Brigitte Kaufmann, Nora Geiser, Alexander Paul Leff

## Abstract

Statistical parametric mapping (SPM) software was implemented in the early 1990s so that neuroscientists could test spatially extended hypotheses using functional imaging data, usually in 3D space and allowing for a mass univariate approach to hypothesis testing that is agnostic to where any significant effects may lie. Here, we apply the same approach to gaze duration data, i.e. visual fixations, collected using a virtual reality headset, which extends across a large 2D area of visual space, measuring 32° either side of central fixation and 24° above and below this point.

In order to evaluate this novel method, we measured the locus of average gaze in a group of 17 patients with hemispatial inattention to the left, a neurological condition caused by damage to the right parieto-frontal brain networks, that induces a systematic bias in lateralised visual attention. This causes people to experience difficulty in paying attention to one side of space, both in their extrapersonal world and relative to their own bodies. We used a free visual exploration paradigm (viewing multiple naturalistic scenes for 7 seconds), which is sensitive to spatial biases encountered in this condition. 23 age-matched and neurologically healthy controls also took part. The visual stimuli were original and mirror flipped versions (Left to Right ie L-R) to correct for any lateralised informational biases inherent in the images. When compared with age-matched controls, the patients exhibited an average spatial shift of attention of 18° to the right of the midline.

We demonstrated this approach using patients with hemispatial inattention, but it can be applied to any fixation-based or dwell time data. This is an advance on current methods that generated visual heatmaps or attentional maps, as our technique allows formal testing of spatially extended hypotheses on gaze duration data using a standard, frequentist statistical approach.

## Introduction

Hemispatial inattention, also known as visual neglect, is a neurological disorder that occurs in 25-30% of stroke sufferers requiring hospitalization in the acute phase (1). It is characterised by a pathological bias in spatial attention, such that patients fail to orient, respond, or explore stimuli presented on the side of space contralateral to the lesion. There is commonly a spatial bias in gaze preference to the right hand side, causing left sided visual neglect, owing to the unilateral processing of left space by the right cerebral hemisphere, although hemispatial inattention has also been described following left hemispheric pathology (2, 3). In clinical practice, this can have major consequences for safety, functional independence, engagement with rehabilitation, overall disability and morbidity outcomes, and length of inpatient stay in hospital (4, 5). Accurate identification and quantification of spatial bias are therefore important both diagnostically and for monitoring recovery or treatment response.

Assessment of hemispatial inattention remains challenging. Conventional approaches include paper-and-pen tests such as cancellation and line bisection tasks, as well as behavioural scales including the Catherine Bergego Scale. However, no single assessment has emerged as a universal gold standard, and different tests can vary in their sensitivity depending on the neglect subtype, spatial domain, and task demands (6, 7). In particular, conventional bedside tools may under-detect impairments affecting extrapersonal exploration, may not fully capture the spatial distribution of exploratory bias, and can show between-test variability (8). These limitations have motivated interest in more sensitive and behaviourally rich methods of quantifying visuospatial exploration.

Eye tracking during free visual exploration offers one such approach. Using eye movement tracking to better understand human cognitive processes has been in use for over a hundred years, but it is the advent of real-time video-oculography that has led to an explosion in clinical and research applications over the last few decades. Modern video-oculography allows detailed measurement of gaze behaviour while participants view naturalistic stimuli, generating high-density spatial and temporal data. Commonly derived measures include saccades (ballistic eye movements that relocate the eyes to a new target in the environment), fixations (the time spent on a certain target between saccades) (9) and dwell times (a summation of the time spent within the co-ordinates marking a single area of interest) (10), and are often represented as scanpaths (usually denoted by lines connecting subsequent gaze fixations) (11), gaze plots (represented by circles marking the position of a fixation, with the radii of the circles signifying the time spent at each fixation) (12), and heat maps (which aggregate the relative frequency of fixations) (13, 14). These representations have proved useful for visualising how observers distribute their attention across scenes, however, to date, there has been no method for producing spatially-extended statistical maps of eye movement data. Indeed, eye-tracking frequency can be recorded at 50 Hz up to 2000Hz with some trackers, leading to large datasets for even relatively short experiments.

Prior work has suggested that eye tracking during free visual exploration may detect neglect-related spatial bias more sensitively than some conventional paper-and-pen measures (15), but methods for the statistical analysis of spatially rich data remain limited. Existing approaches to eye-movement analysis provide several valuable ways of characterising visual exploration, but also have their limitations. For example, scanpath-comparison methods can capture the temporal sequence and organisation of gaze shifts across a scene, while saliency-map and heatmap approaches provide useful summaries of how attention is distributed across image space and how fixation behaviour relates to visual scene structure, including attempts to predict fixation selection from image salience (16, 17). These methods have important strengths, but they are not primarily designed to support voxel-wise or grid-wise statistical inference across spatially extended gaze data. In particular, heatmaps and related visualisations are often descriptive, and many approaches either rely on pre-specified regions of interest or reduce gaze behaviour to a limited number of summary indices. As a result, they may not fully preserve the spatial richness of the original data or provide a principled framework for testing where reliable group differences occur across the visual field. The novel approach we present addresses this by applying spatially distributed statistical modelling to gaze-duration data, allowing formal inference across the explored field while preserving the dimensional richness of the data.

Statistical Parametric Mapping (SPM) provides such a framework (18). Originally developed for functional imaging data from neuroimaging, SPM software has been used since the early 1990s, enabling mass-univariate statistical testing across spatially organised data, allowing valid inferences to be made at each location within an image while controlling for multiple comparisons across the full search space. Rather than averaging over pre-defined regions of interest, it allowed neuroscientists to ask where in a spatial field a significant effect occured. In functional neuroimaging, this logic is typically applied to three-dimensional brain volumes made up of many thousands of identically shaped cubic elements (i.e. voxels). Conceptually, however, the same approach can be extended to other forms of spatially structured data. In the present context, our eye-tracking data can be represented as two-dimensional maps of viewing time across the visual field. We can therefore extend the statistical approach of SPM to this spatially extended gaze behaviour data. This consists of applying linear regression at each pixel of the images being analysed, to yield a statistical map (18). A correction for multiple comparisons across pixels is then applied, using an approach that derives from the field of topology, called Random Field Theory (19).

This approach thereby permits the use of first-level models to estimate image-specific or within-subject effects and second-level models to test group effects across participants. The principal advantage of this method is that it preserves the spatial richness of the data while enabling formal statistical inference across the entire explored field, without restricting analysis to pre-defined regions or reducing the data to a single summary score. The statistical foundations and software for performing these analyses are well established and thoroughly validated, but have never been applied to gaze duration and location maps, until now.

We demonstrated this method in patients with hemispatial inattention after acute stroke, a clinical population in whom lateralised gaze bias is both theoretically relevant and clinically important, comparing them with age- and sex-matched healthy controls. Free visual exploration of naturalistic scenes is well suited to this purpose because it captures spontaneous allocation of attention under conditions that are closer to everyday exploration than many conventional structured tests (20, 21). We hypothesised that statistical parametric mapping of gaze duration would detect and localise spatially distributed differences between patients with hemispatial inattention and healthy controls during scene viewing, thereby providing a principled method for analysing eye-movement data as spatially extended statistical maps.

## Methods

### Participants

Two groups of participants were recruited (Table 1). The patient group comprised 17 people with stroke who were still inpatients, having been admitted either to an Acute Stroke Unit or a Neuro-Rehabilitation Unit, and had been identified by the multidisciplinary teams as having hemispatial inattention. The patient group was 53% female and had a mean age of 62.46 years (SD 12.33). The control group comprised 23 neurologically healthy participants with no previous history of stroke or ophthalmological disease. The control group was 65% female and had a mean age of 68.96 years (SD 9.56). The groups were matched for age and sex; an independent-samples t-test showed no significant age difference, t(38) = 1.82, p = 0.08, and a chi-square test showed no significant sex difference, χ^2^(38) = 6.14, p = 0.433. Recruitment began on 24 November 2022 and ended on 30 March 2023. Data from the patient group were collected on the inpatient wards, while healthy controls were recruited locally through the University. We were able to obtain MRI brain scans for 16/17 patients. Using SPM we created a lesion overlap map to display the extent and commonalities of their lesion topography (24). The greatest intensity (red, Figure 1) was in the Right Middle Cerebral Artery territory, which includes the right parietal lobe and its frontal connections, areas most commonly implicated in people with left-sided hemispatial inattention (25).

**Table 1:** Star Cancellation values, laterality indices, Broken Hearts Asymmetry scores, Catherine Bergego Scale scores and the x-coordinate for the centre of gaze (minimum being 0 from the left to maximum 32 towards the right) for patients. Bold values are those in the diagnostic range for lateralised inattention.

| *Stroke Participants* |  |  |  |  |  |  |  |  |  |
| --- | --- | --- | --- | --- | --- | --- | --- | --- | --- |
| ID | Age | Gender | Total Stars<br>Baseline | Stars<br>cancelled<br>-Left | Stars<br>cancelled<br>-Right | Star<br>Cancellation<br>Laterality<br>Index | CBS<br>Baseline | Broken<br>Hearts<br>Space<br>Asymmetry | X-coordinate<br>for centre<br>of gaze |
| CT01 | 50-54 | F | 43 | 19 | 24 | **0.44** | 9 | - | 31 |
| CT02 | 60-64 | M | **27** | 1 | 26 | **0.03** | **15** | **12** | **26** |
| CT03 | 50-54 | M | **22** | 5 | 17 | **0.22** | **16** | **11** | **23** |
| CT05 | 45-49 | F | **31** | 15 | 16 | **0.48** | **20** | **11** | **20** |
| CT07 | 70-74 | F | **33** | 7 | 26 | **0.21** | **14** | **9** | **22** |
| CT08 | 55-59 | F | 45 | 20 | 25 | **0.44** | **17** | 0 | 15 |
| CT09 | 65-69 | F | 49 | 24 | 25 | 0.48 | **14** | **6** | **23** |
| CT11 | 60-64 | M | **20** | 1 | 19 | **0.05** | 9 | **13** | **24** |
| CT12 | 70-74 | M | **20** | 0 | 20 | **0** | **12.5** | **15** | **20** |
| CT13 | 60-64 | M | **6** | 0 | 6 | **0** | **14.4** | **22** | **22** |
| CT14 | 70-74 | M | **37** | 10 | 27 | **0.27** | **26** | **7** | **24** |
| CT15 | 75-79 | F | **15** | 0 | 15 | **0** | **11.25** | **4** | **27** |
| CT16 | 80-84 | M | **32** | 13 | 19 | **0.40** | 6 | **15** | **19** |
| CT17 | 50-54 | F | **25** | 4 | 21 | **0.16** | 9 | **15** | **21** |
| CT18 | 30-34 | F | **7** | 0 | 7 | **0** | **17** | **11** | **28** |
| CT19 | 65-69 | M | **13** | 0 | 13 | **0** | **14** | **19** | **28** |
| CT21 | 65-69 | F | **8** | 0 | 8 | **0** | **27** | **20** | **20** |

**Figure 1.**
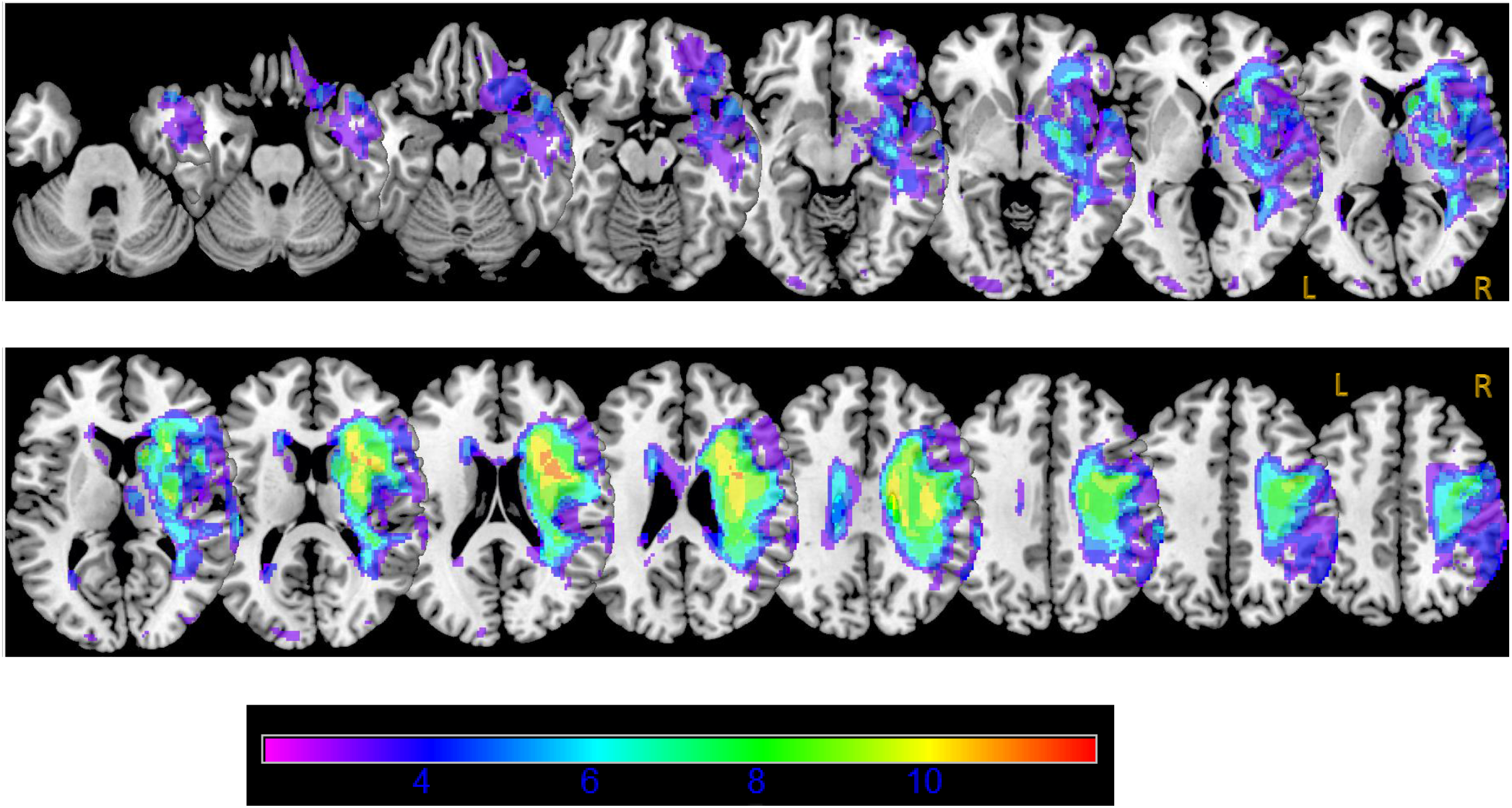
Lesion Overlap Map. The patient’s lesions have been displayed on a canonical MRI T1 weighted image in standard MNI space. Axial slices in ascending steps of 2,3 or 5mm are oriented in neurological convention (right side of the brain on the right of the images - denoted by R and L on the final slices as an example). The colour intensity scale indicates the number of patients who have a lesion affecting each voxel.

### Clinical Assessment of Hemispatial Inattention

The presence and severity of hemispatial inattention were assessed using clinical examination, two impairment-based cancellation tests, and one functional measure. The impairment-based measures were the Star Cancellation Test and the Broken Hearts Test from the Oxford Cognitive Screen, while the functional measure was the Catherine Bergego Scale (CBS), scored by a treating therapist. Cut-offs were used to screen in patients with at least a moderate degree of hemispatial inattention. Moderate severity was defined as a Star Cancellation score of fewer than 42 out of 54 stars together with a laterality index between 0 and 0.46, a Broken Hearts space asymmetry score between 4 and 12, or a CBS score of at least 11 out of 30. In the patient cohort, the mean Star Cancellation score was 25.4 (SD 12.99), the mean Star Cancellation laterality index was 0.18 (SD 0.19), the mean Broken Hearts space asymmetry score was 11.87 (SD 5.7), and the mean CBS score was 14 (SD 4.69). One patient met diagnostic threshold on one of these four criteria, two met threshold on two criteria, three met threshold on three criteria, and the remaining 11 met all four. Control participants performed at ceiling on the two cancellation tests.

### Ethics and Consent

All participants were provided with an information sheet describing the use of a virtual reality headset with eye tracking to capture gaze behaviour, and written informed consent was obtained. For the patient group, capacity had already been assessed by the referring clinical team as part of their eligibility assessment for participation in the standardised neuro-rehabilitation programme, and this information was reviewed by the research team prior to consent. Healthy controls were not formally assessed for capacity. In both groups, participants were given a 24-hour period to review the information sheet, comprehension of participation was confirmed, and any questions were addressed before consent. Ethical approval was granted by the UCL Research Ethics Committee (Project ID 22941/001).

### Virtual Reality: Stimulus Presentation

The free visual exploration task used naturalistic scene stimuli presented as landscape-formatted 2D images which were sourced from Kaufmann et al.(30) (Figure 2). They were imported into custom written software so that they could be displayed in the HTC Vive virtual reality headset. There were two main advantages to importing these images into the Virtual reality headset: firstly, the images were made to subtend a greater visual angle in the Vive (64° by 48°, as opposed to 28° by 21° in the original version(20)); secondly, eye movements were measured using the built-in Vive eye-tracker which corrected for any head movements. The 24 images comprised of 12 pairs of original images and their laterally flipped mirror images. This approach was used to minimise intrinsic image-driven lateralised bias arising from visual saliency.

**Figure 2:**
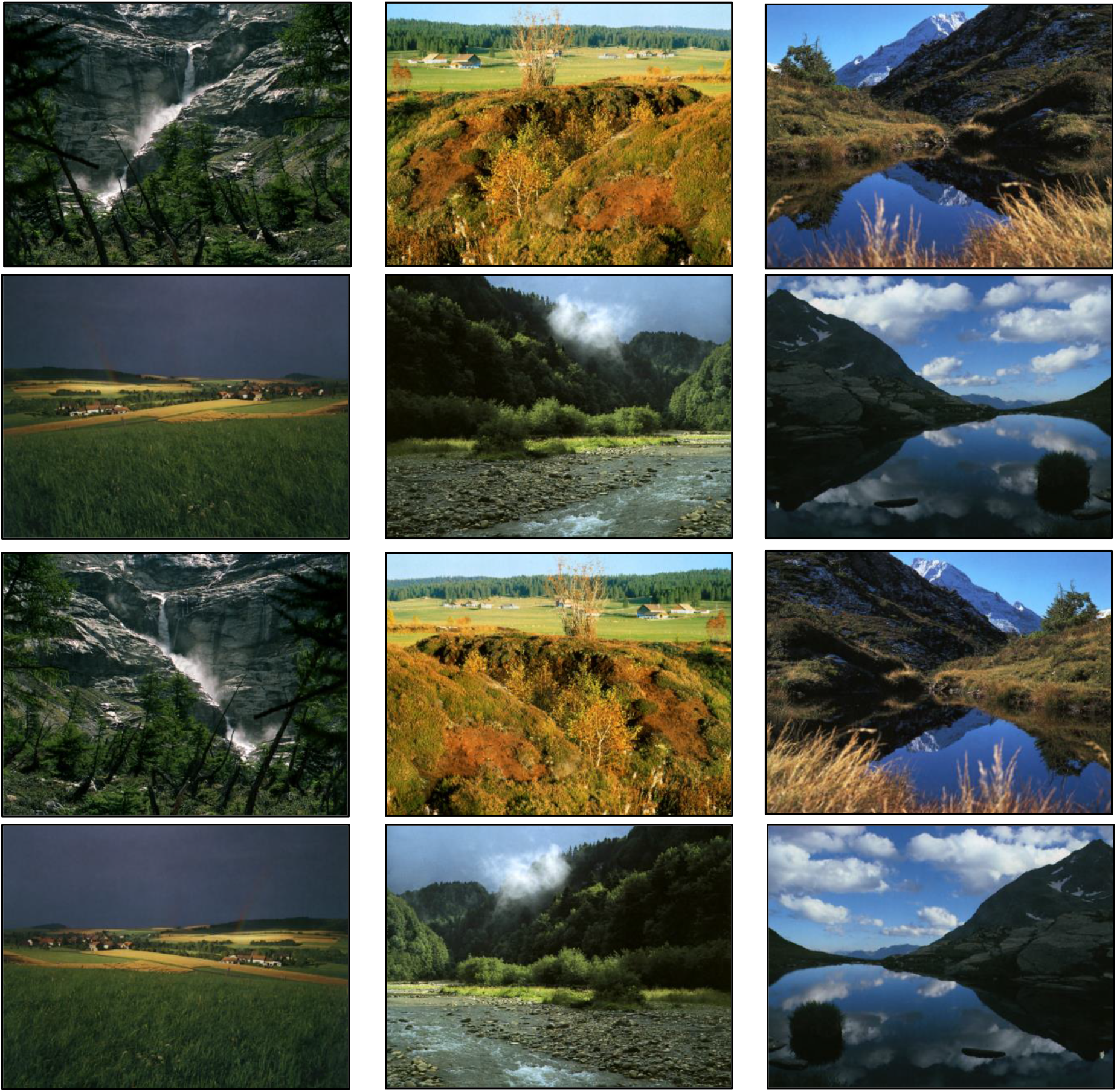
Six pairs of original images and their reflected mirror images displayed within the Vive headset. They are presented here as they would appear to a participant, with no original image and mirror image pair appearing consecutively, and no image being repeated.

### Apparatus and Setup

Stimuli were presented using a head-mounted virtual reality headset incorporating eye tracking. An HTC Vive Pro Eye Headset, which has eye-tracking and room-scale tracking via the use of positional tracking base stations, was used to capture gaze fixation data. The HTC Vive Pro Eye Headset utilizes Tobii technology to power its eye-tracking capabilities. SteamVR, developed by Valve, was used as the software platform for headset setup and stimulus presentation. Prior to testing, the headset was fitted and calibrated via the SteamVR dashboard to align the visual display and eye-tracking system with the participant’s midline and viewing height while they were positioned either in an inpatient bed or seated in a chair. This calibration was performed to optimise comfort, ensure stable visual alignment, and improve registration of gaze with the displayed stimuli. Images remained centrally fixed within the headset irrespective of head movements. The software was operated on an MSI GT73VR GRF Titan Pro laptop, equipped with an Intel Core i7-6700HD @ 2.60HZ processor, 16GB dual-channel DDR4 RAM, NVIDIA GeForce GTX1080 graphics card, running on a 64-bit version of Windows 10 Home.

### Protocol

Participants were instructed simply to view the images, without giving any verbal description or feedback. Original images and their corresponding mirror images were presented as separate stimuli in a pseudorandom order such that no original image and its mirrored counterpart appeared consecutively. Each image was displayed for 7 seconds. The inter-stimulus interval was 3 seconds and consisted of a white fixation cross on a grey background displayed at the centre of the field of view. The total task duration was approximately 4 minutes.

### Eye Movement Data

The Tobii technology within the HTC Vive Pro Eye Headset powered its eye-tracking capabilities. Within the headset, illuminators shine near-infrared light on to the user’s eyes, creating reflections which are captured in high-resolution by built-in cameras. This information is fed into image processing algorithms that extract data relating to a variety of outputs such as gaze origin, gaze direction, pupil position, pupil size and eye openness. The headset has a 110° trackable field of view, with binocular gaze data sampling frequency of 120Hz, and an accuracy of 0.5°–1.1° (within a 20° field of vision). During blinking, the eyelids close thereby obstructing any pupillary or corneal reflections of near-infrared light, and therefore these periods do not register as data points.

In the case of our study, using custom-written software, gaze location and duration data for each image viewed in the Vive Pro Eye Headset was outputted into a Microsoft Excel Comma Separated Values (.csv) file. Each Excel .csv file can be thought of as representing a two-dimensional grid over the stimulus, comprising 24 × 32 grid cells. Each grid cell subtended approximately 2° of visual angle, corresponding to a total explored visual field of 32° to either side of the horizontal midpoint (64° total horizontally) and 24° above and below the vertical midpoint (48° total vertically). Each grid cell in which gaze was maintained for more than 100 ms was assigned a value corresponding to the cumulative gaze duration for that location, rounded to the nearest 100 ms. For example, if the gaze fixation time on a particular grid cell was 220ms, this would generate a value of “2” in that grid cell. The 100ms threshold was used to preferentially capture voluntary fixations and to exclude very brief transient gaze samples. The 24 × 32 grid represented a pragmatic trade-off between spatial precision and reliable registration of gaze across neighbouring cells; smaller cells risked sparse or missed registration as gaze moved across the display, whereas larger cells would have reduced spatial resolution. Grid size was not systematically varied in the present study, and its influence on results should be assessed in future work. Dwell time was cumulative so could be generated by one or more fixations. The data was outputted in an excel spreadsheet with integer values for each grid cell, one sheet for each image viewed. Eye-tracking analyses were conducted on raw gaze location and duration data; no separate fixation extraction procedure was applied.

### SPM Pipeline and Analysis

#### (i) Pre-Processing

Each Excel spreadsheet (one for each image viewed), was then converted into a NIfTI (.nii) file (Figure 3), to allow for further analysis within SPM.

**Figure 3:**
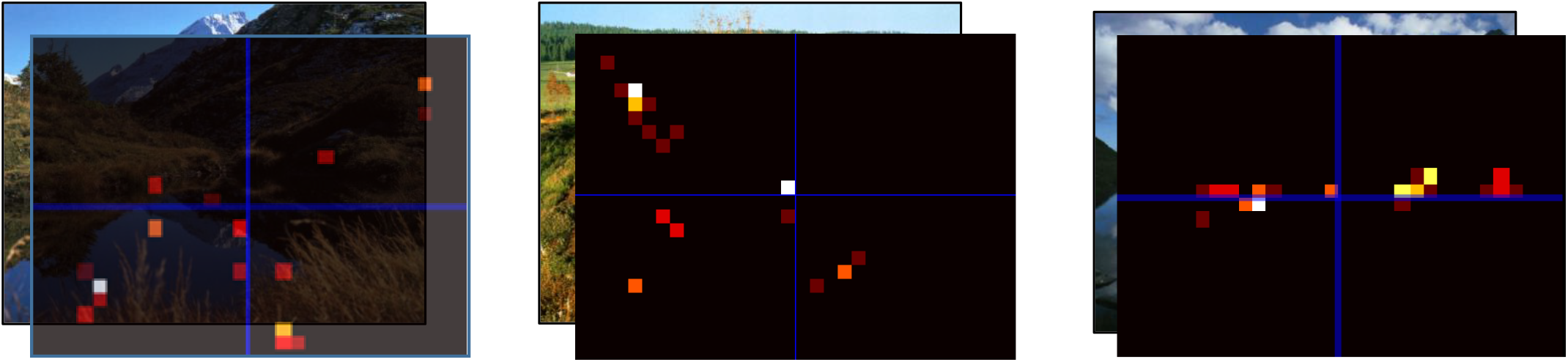
The first image on the left with the semi-transparent raw heatmap demonstrates the super-imposition of converted .nii files representing the 24 x 32 cells grid over the image underlying it, with each coloured point representing gaze fixation over that cell for >100milliseconds. These **raw** heatmaps were generated for each image, as can be seen in the next two images. The colour gradient from red to orange to yellow to white reflects duration of dwell time, starting with a minimum of 100ms (red). The blue crosshair is at the centre of the heatmap.

Each .nii file was smoothed using a Gaussian smoothing kernel with 8mm full-width half maximum in the pre-processing step in order to increase the signal to noise ratio (Figure 4i,ii).

**Figure 4 (i,ii, A –H):**
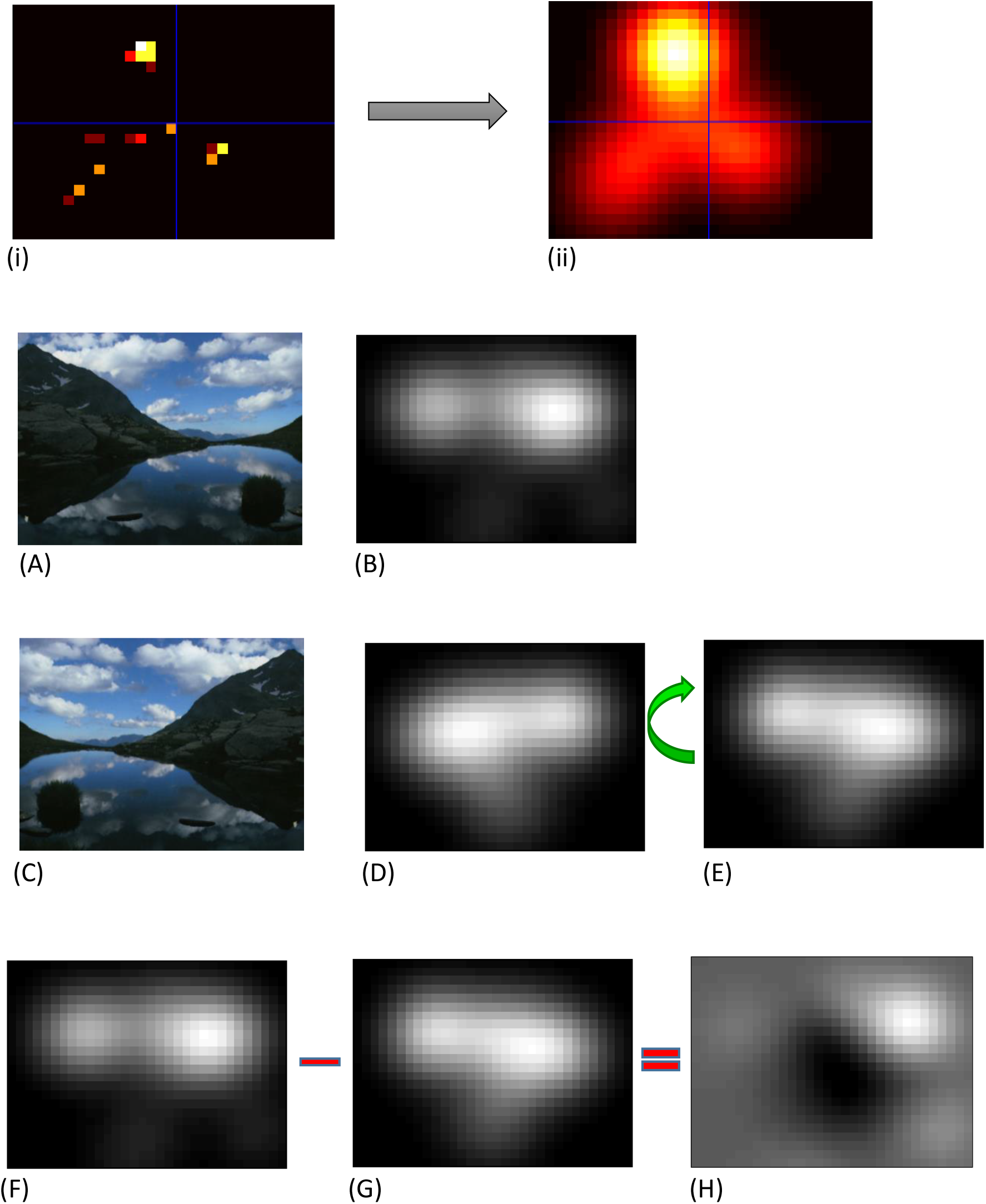
i to ii shows the .nii file smoothed using an 8mm scale in the pre-processing stage (A) An example of an Original Image and (B) the resultant smoothed heatmap (greyscale) of dwell times from a single subject viewing the image for 7 seconds. (C) The Mirror Image of the original (D) with corresponding smoothed dwell time heatmap. (E) The Flipped version of the Mirror Image smoothed heatmap.(F-H) Subtracting the Flipped Mirror Image heatmap [G] from the Original Image heatmap [F] (with the Saliency points aligned and therefore cancelling each other out) to produce the “Original Minus Flipped Mirror Image” [H] for each pair.

For each participant, the 24 .nii files were then separated into 2 groups of twelve, titled the ‘Original Image’ group which included the .nii files of 12 unique images, and the ‘Mirror Image’ group, which included the respective .nii files for the corresponding Mirror images for each Original image.

#### (ii) Negating any horizontally-expressed salient features

Our aim was to characterise and quantify the degree of lateralised bias in the patient group while viewing naturalistic images, but in order to do this we need to address the issue saliency inherent in such images. Salient image features are areas of perceptual prominence that garner more attention, and therefore would be expected to have more local fixations and thus dwell times(23). The study of gaze fixation density maps, which reflect the likelihood of pixels in an image being viewed by human observers, reveals that salient areas on an image tend to be those that are rich with structural information, and that each image will only have a limited area which has a high saliency value. Visual saliency had to be specifically considered in this work as we are using images and video-oculography to assess the presence of spatial bias. Salient features (also termed “saliency points”) on an image are defined as areas of perceptual prominence that garner more attention, and therefore would be expected to have more fixations, with initiation of saccades and shifts of attention towards them(23).

In order to control for intrinsic, saliency-led lateralised bias in the 12 images we used to generate the 2D dwell maps, we created a laterally flipped or ‘mirror’ image for each one. Therefore, all the smoothed ‘Mirror Image’ dwell time maps were flipped laterally (therefore along the vertical axis), using the Image Calculator feature within SPM, with the expression *flip(i1,1)*. This enabled us to then proceed to the next step, during which we subtracted the Flipped Mirror Image from the Original Image, using once again the Image Calculator feature, with the expression *(i1-i2)* (Figure 4A-H). These steps provide a 2D average dwell time map for any given image with horizontally-expressed salience features factored out. This step in the methods was incorporated in order to negate any horizontally-expressed salient features.

A subject viewing both scenes by fixating on exactly the same features would produce a null subtraction image. A subject who viewed both images with a lateralised spatial attentional bias would have areas with high positive values (to the side where gaze was preferentially directed, i.e. in our study towards the right side) with high negative values towards the neglected side.

### 1^st^ Level Analysis: Within-Subject

For each subject, a statistical map was generated by performing a linear regression at each pixel of the image using the standard tools in SPM. For the pixel with index *i*, the General Linear Model (GLM) was specified:

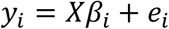

Where *y*_*i*_ is a vector of values from the 12 subtracted images for the *i*-th pixel and *X* is the *design matrix*, the columns of which are hypothesised effects (a.k.a covariates, regressors or explanatory variables). For this example the design matrix consisted only of a column of ones, in order to calculate the average of the images. Residuals *e*_*i*_ were estimated using the standard Restricted Maximum Likelihood (REML) scheme implemented in the SPM software. The output of this stage of the analysis was an image of regression parameters for each subject.

### 2^**nd**^ Level Analysis: Between-subjects (people with lateralised inattention vs. control subjects)

The 23 regression parameter images from the 1^st^ level analysis for the control subjects, were compared against the 17 parameter images for the patients. This was implemented by specifying a design matrix consisting of a dummy variable for each group (Figure 5A). In terms of modelling variance, the groups were treated as independent with unequal variance, meaning that any potential heteroscedasticity was modelled by estimating separate covariance components for each group. An F-contrast of [1, -1] was used to test for a difference in lateralised gaze dwell times across the two groups (top part of Figure 5A). The p-value threshold was set to 0.05 with a Family Wise Error rate calculated using random field theory(19).

**Figure 5A&B.**
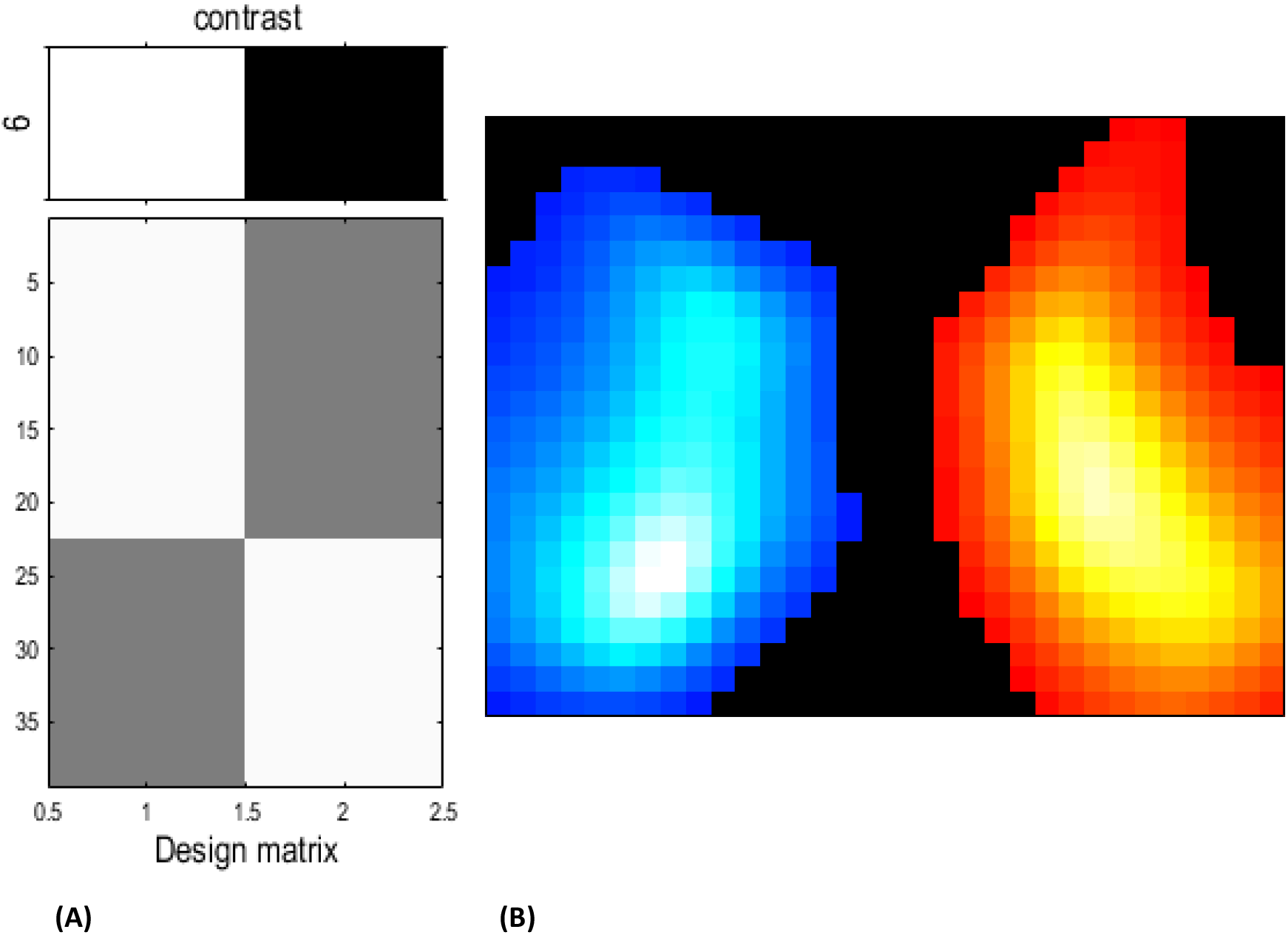
The Design Matrix on the left (A) displays the schematic of the f contrast of 1 -1 applied to the 2 groups, with controls’ images entered into the left column and patients’ on the right. The SPM on the right (B) shows the two areas where patients’ gaze dwelt significantly more (warm colours) and less (cool colours) than the control subjects.

### Exploratory correlation analysis

To explore the relationship between the free visual exploration gaze metric and established clinical measures of neglect severity, exploratory correlation analyses were performed between the x-coordinate of the centre of gaze during free visual exploration and three bedside measures: total stars missed on the Star Cancellation Test, left-sided stars missed on the Star Cancellation Test, and Catherine Bergego Scale score. Higher x-coordinate values reflected a more rightward centre of gaze and therefore greater left-sided neglect. Distributional inspection using histograms and Shapiro–Wilk tests showed that the x-coordinate variable, total stars missed, and CBS were not significantly non-normal, whereas left-sided stars missed showed a markedly skewed distribution with clustering toward the upper end. Given the modest sample size and this non-normality, non-parametric Spearman correlations were used.

## Results

The group results of the comparison between control subjects and patients are displayed in Figure 5B, thresholded at p <0.05 FWE corrected. This SPM is in image space measuring 64° by 48° of visual angle. Two areas of significant differences were identified, a cluster of 256 voxels to the right side of space where the patients had higher dwell times than the controls, and a cluster of 271 voxels to the left, where they had lower dwell times (Fig 5B and Table 2).

**Table 2.**
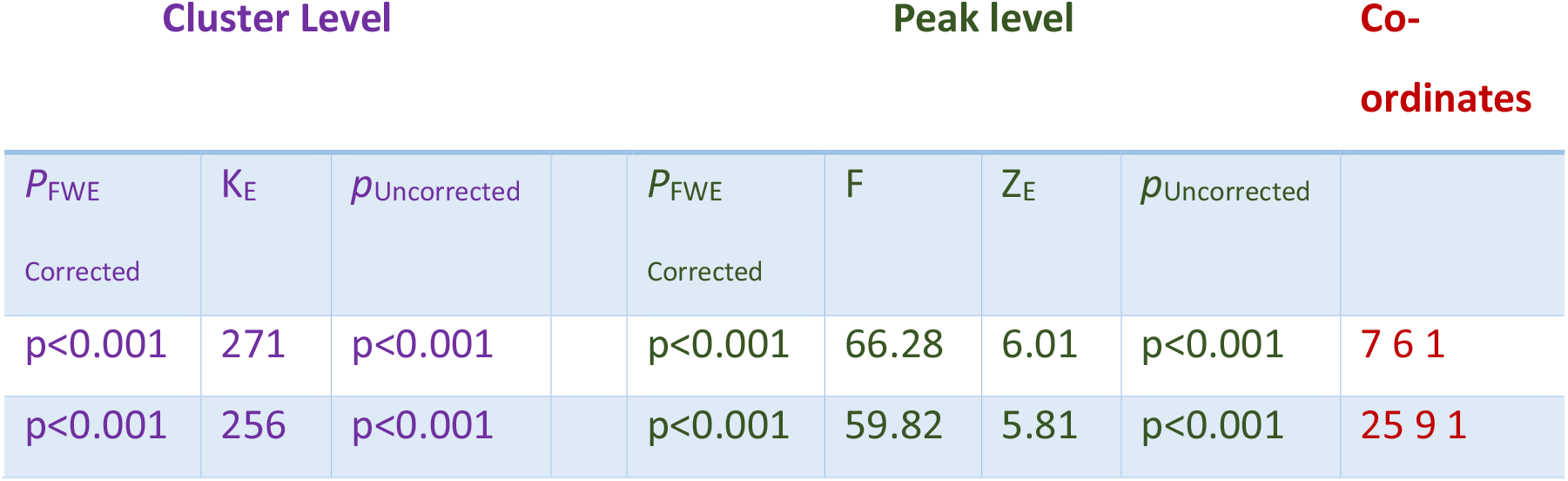
Cluster level (left side) and peak voxel (right side) significance levels from the SPM in Fig 6. P_FWE_ is the p value corrected using Family Wise Error correction for multiple comparisons. KE is the size of the cluster in contiguous voxels. The co-ordinates are in the image space and give the location of the two peaks of difference in the SPM. The bottom left voxel of the SPM has the co-ordinate of 0 0 1 (x, y, z). The SPM is 2D so has a thickness (z) of 1 throughout.

The peak voxel for the patient group is highlighted in Figure 6 with the data from the two groups at this point extracted and plotted as a bar graph (Graph 1).

**Graph 1:**
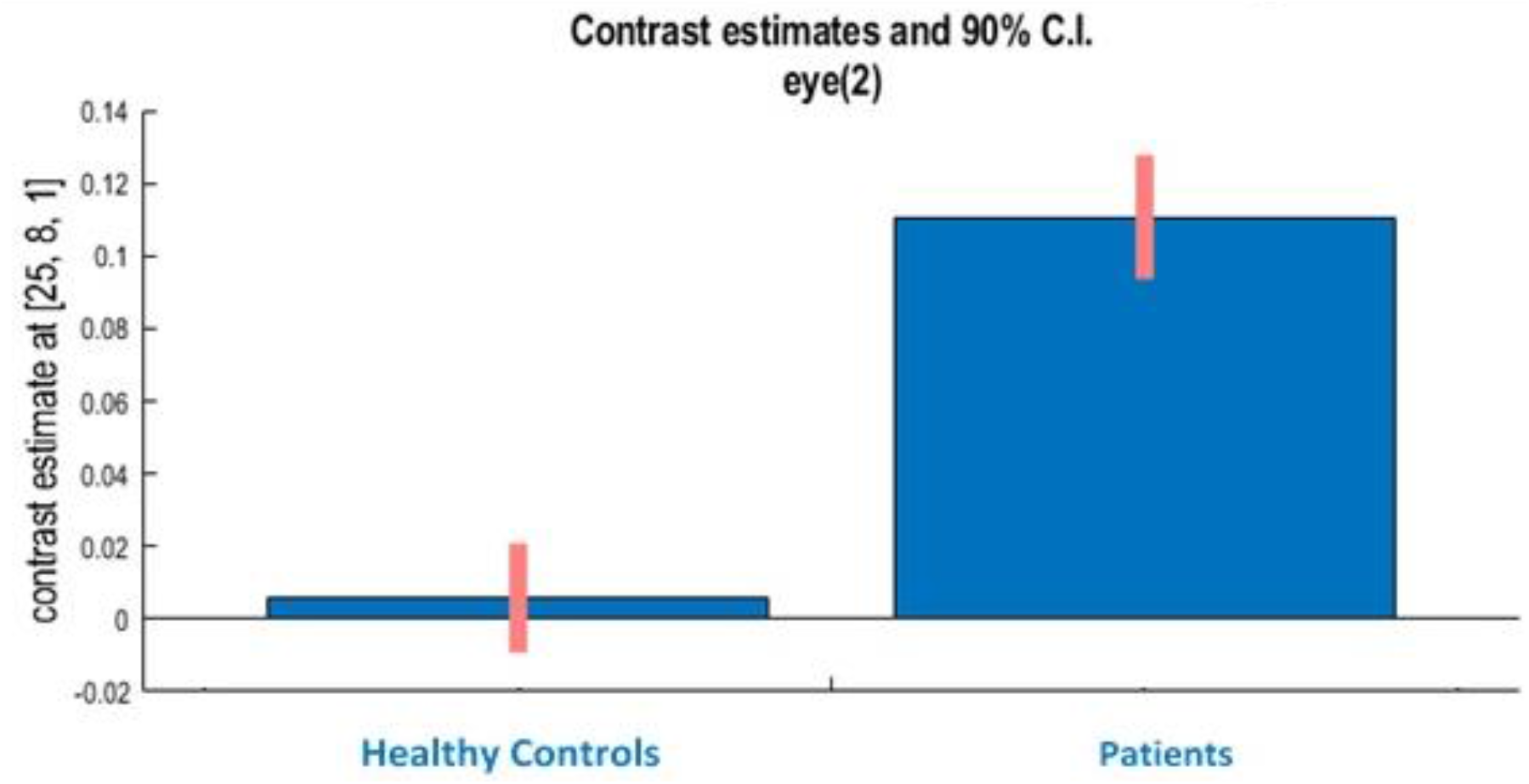
The peak voxel in Figure 6 with data from the Patient group and the Healthy controls extracted and plotted as a bar graph.

**Figure 6:**
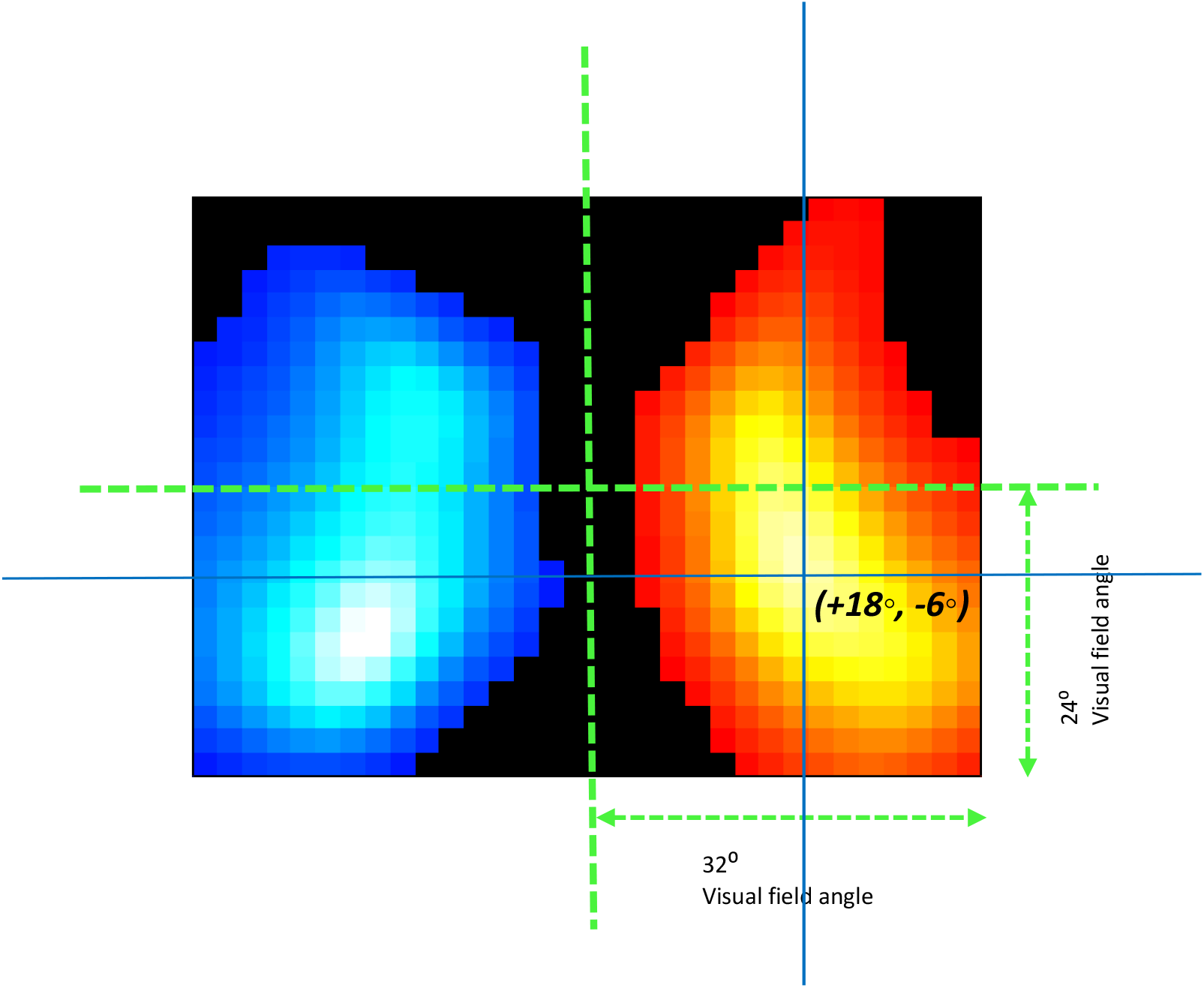
On the left, the SPM from Fig 5 is reproduced but with the centre of the image marked with green cross hairs. The peak average centre of gaze for the patient group (blue crosshairs) deviated from the midline by 18 degrees over to the right of centre, and six degrees below the horizontal meridian. Data from the peak voxel is plotted on the right (NB: Y-axis is in arbitrary units).

Exploratory Spearman correlation analyses were performed to assess the relationship between the x-coordinate of the centre of gaze during free visual exploration and clinical neglect measures. No significant correlations were observed with total stars missed on the Star Cancellation Test (rho = 0.184, p = 0.479), left-sided stars missed (rho = 0.246, p = 0.342), or Catherine Bergego Scale score (rho = -0.126, p = 0.631).

## Discussion

The advent of video-oculography has brought new opportunities for the assessment of gaze location and duration, but also poses challenges for statistical analysis if one wishes to preserve the richness of the spatial data in the co-ordinate space in which it was collected. We have described, for the first time, a method for carrying out statistical analyses on spatially extended gaze location and duration data using statistical parametric mapping. In the present study, this approach detected a marked rightward spatial bias in patients with hemispatial inattention relative to healthy controls during free visual exploration of naturalistic scenes. Taken together, these findings provide a proof-of-principle that inferential methods such as SPM, originally developed for neuroimaging, can indeed be adapted to eye-movement data in order to detect and localise pathological spatial bias.

We have used patients with visuospatial inattention as an example because they have an inherent, lateralized spatial gaze bias, but this approach can be used for any fixation-based or dwell time eye-movement related data. It can be used on groups, as we have done here, but it can also be carried out on single subjects, if they view enough stimuli A key contribution of the present method is that it complements existing eye-tracking outputs such as scanpaths, gaze plots, and heatmaps by enabling formal statistical inference across spatially extended gaze data.This is different from previous methods that have generated visual heatmaps or attentional maps, as our technique allows formal testing of spatially extended hypotheses on gaze location and duration data using a standard, frequentist statistical approach. Heatmaps and attentional maps remain useful descriptive tools, but they are often primarily visual summaries of aggregated gaze behaviour rather than frameworks for corrected inferential testing across the full explored field. The current approach therefore offers a more principled way of examining spatially distributed gaze abnormalities without collapsing the data into a small number of summary measures.

These findings should also be considered in the broader context of neglect assessment. There is no single gold-standard test for hemispatial neglect, and different bedside or behavioural measures capture different aspects of the syndrome, with variability in sensitivity depending on spatial domain, task demands, and neglect subtype (7, 8). Conventional paper-and-pen measures remain clinically useful, but they may under-detect impairments outside peripersonal space and do not necessarily capture the full spatial distribution of exploratory bias. In this context, free visual exploration is attractive because it samples spontaneous visual-attentional behaviour under relatively naturalistic conditions. Prior work by Kaufmann and colleagues does support the view that video-oculography has proven to be more sensitive in detecting visual inattention than conventional paper-based tests (15).

We performed additional exploratory correlations between the x-coordinate of the centre of gaze during free visual exploration and conventional neglect measures, namely total stars missed, left-sided stars missed, and Catherine Bergego Scale score. These analyses did not demonstrate significant associations in the present sample. This may reflect limited statistical power, restricted range or skewness in some bedside measures, or the possibility that a single centre-of-gaze summary metric does not fully capture the spatially extended abnormalities targeted by the present SPM-based approach. In particular, left-sided stars missed showed a clearly skewed distribution with clustering toward the upper end, and for this reason non-parametric Spearman correlations were used. The absence of significant correlations therefore does not necessarily imply that the present method lacks clinical relevance, but rather that the relationship between spatially extended gaze-derived measures and conventional bedside assessments may not be one-to-one and requires further validation in larger cohorts.

Our current approach does have practical advantages to consider. Eye-tracking within Virtual Reality headsets carries the advantage of an increased field of view over more traditional 2D displays. In the case of our study, whilst the HTC Vive Headset allows up to 110° field of view, the 2D images that we uploaded allowed us to measure 64 x 48 degrees of visual angle, enabling us to detect the location of spatial bias more accurately. Kaufmann et al., who also utilized a free visual exploration task by displaying their images on a large monitor(30), were only able to test a maximum of 32° of visual angle along the horizontal plane. Our technique doubles this along both visual planes, allowing us to sample four times the visual area. This theoretically makes the test more reliable in terms of detecting the extent of any spatial bias. The average displacement in our study was 18° to the right of centre. The standard pen-and-paper test of hemispatial inattention, the Star Cancellation Test on an A4 sheet of paper, when viewed at 50 centimetres distance, only subtends a maximum angle of 15° to either side. Overall, the images in the HTC Vive Headset subtended a substantially larger visual angle than in the original monitor-based implementation, allowing a broader region of visual space to be sampled during exploration. This may be advantageous when the aim is to characterise extrapersonal spatial bias more fully than is possible with standard bedside tests. At the same time, the present approach is analytically more complex than conventional paper-and-pen assessment and also more demanding than simpler gaze summary measures such as mean gaze position alone.

By using flipped versions of each naturalistic scene, we were able to correct the gaze data for any inherent lateralized spatial biases in the images. We did not do this for the vertical components and this probably explains why we identified a significant vertical displacement of 6° below the horizontal meridian. This is in the opposite direction from what one might expect. A vertical neglect component (albeit of smaller magnitude compared to that seen in the horizontal plane) has been described previously in people with hemispatial inattention, with a bias appearing in the upper quadrants (32, 33). The naturalistic images used in our free visual exploration task generally contained more areas of interest and higher salience in the lower quadrants as many were outdoor scenes comprising more featureless sky at the top.

In conclusion, the results generated in this study serve as an archetypal example of a statistical outcome from this novel method that allows for the statistical analysis of spatially distributed gaze data, regardless of the visual capture method utilized. This approach to eye-movement behaviours in response to visual stimuli offers numerous applications across a variety of disciplines, such as task-based visual assessments like driving, neuropsychological and neurobehavioural gaze assessments relevant to industries like advertising, the arts and cognitive neuroscience. The method successfully applies statistical parametric software on to a challenging data set, at a time that is particularly relevant given the rise of advanced video-oculography and eye-tracking data now available from online testing platforms, and extends beyond descriptive visualisation by enabling formal analysis of spatially distributed gaze behaviour.

A number of limitations should be acknowledged. The sample size was modest, the additional clinical correlation analyses were exploratory, and the gaze validation metric used here (the x-coordinate for the centre-of-gaze) may not fully capture the spatially extended abnormalities that the SPM-based approach is designed to detect, and explain the lack of correlation with existing measures. In addition, methodological choices such as grid size and duration threshold were selected pragmatically and were not systematically varied in the present study. Future work should therefore evaluate this approach in larger cohorts, test reproducibility across broader datasets, and clarify how this method can best be positioned as an advantageous tool alongside existing clinical and eye-tracking approaches in the assessment of hemispatial neglect.

## Data Availability

All data produced in the present study are available upon reasonable request to the authors

## REFERENCES

1. Ferro JM, Mariano G, Madureira S. Recovery from aphasia and neglect. Cerebrovasc Dis. 1999;9 Suppl 5:6–22.

2. Li K, Malhotra PA. Spatial neglect. Pract Neurol. 2015;15(5):333–9.

3. Stone SP, Halligan PW, Greenwood RJ. The incidence of neglect phenomena and related disorders in patients with an acute right or left hemisphere stroke. Age Ageing. 1993;22(1):46–52.

4. Wee JY, Hopman WM. Comparing consequences of right and left unilateral neglect in a stroke rehabilitation population. Am J Phys Med Rehabil. 2008;87(11):910–20.

5. Chen P, Hreha K, Kong Y, Barrett AM. Impact of spatial neglect on stroke rehabilitation: evidence from the setting of an inpatient rehabilitation facility. Arch Phys Med Rehabil. 2015;96(8):1458–66.

6. Singh NR, Leff AP. Advances in the Rehabilitation of Hemispatial Inattention. Curr Neurol Neurosci Rep. 2023;23(3):33–48.

7. Appelros P, Nydevik I, Karlsson GM, Thorwalls A, Seiger A. Assessing unilateral neglect: shortcomings of standard test methods. Disabil Rehabil. 2003;25(9):473–9.

8. Williams LJ, Kernot J, Hillier SL, Loetscher T. Spatial Neglect Subtypes, Definitions and Assessment Tools: A Scoping Review. Front Neurol. 2021;12:742365.

9. Land MF, Tatler BW, Oxford Scholarship Online P. Looking and acting : vision and eye movements in natural behaviour. Oxford: Oxford University Press; 2009.

10. Vansteenkiste P, Cardon G, Philippaerts R, Lenoir M. Measuring dwell time percentage from head-mounted eye-tracking data--comparison of a frame-by-frame and a fixation-by-fixation analysis. Ergonomics. 2015;58(5):712–21.

11. Dewhurst R, Nystrom M, Jarodzka H, Foulsham T, Johansson R, Holmqvist K. It depends on how you look at it: scanpath comparison in multiple dimensions with MultiMatch, a vector-based approach. Behav Res Methods. 2012;44(4):1079–100.

12. Duchowski A. Aggregate gaze visualization with real-time heatmaps. Eye Tracking Research and Applications Symposium (ETRA). 2012.

13. Stellmach S. 3D Attentional Maps-Aggregated Gaze Visualizations in Three-Dimensional Virtual Environments. In: Nacke LD, Raimund, editor. 2010. p. 345–8.

14. Špakov O. Visualization of eye gaze data using heat maps. ELEKTRONIKA IR ELEKTROTECHNIKA MEDICINE TECHNOLOGY. 2007;T. 115.

15. Delazer M, Sojer M, Ellmerer P, Boehme C, Benke T. Eye-Tracking Provides a Sensitive Measure of Exploration Deficits After Acute Right MCA Stroke. Front Neurol. 2018;9:359.

16. Le Meur O, Baccino T. Methods for comparing scanpaths and saliency maps: strengths and weaknesses. Behav Res Methods. 2013;45(1):251–66.

17. Nuthmann A, Einhauser W, Schutz I. How Well Can Saliency Models Predict Fixation Selection in Scenes Beyond Central Bias? A New Approach to Model Evaluation Using Generalized Linear Mixed Models. Front Hum Neurosci. 2017;11:491.

18. Friston KJ, Holmes AP, Worsley KJ, Poline JP, Frith CD, Frackowiak RSJ. Statistical parametric maps in functional imaging: A general linear approach. Human Brain Mapping. 1994;2(4):189–210.

19. Flandin G, Friston KJ. Analysis of family-wise error rates in statistical parametric mapping using random field theory. Hum Brain Mapp. 2019;40(7):2052–4.

20. Kaufmann BC, Knobel SEJ, Nef T, Muri RM, Cazzoli D, Nyffeler T. Visual Exploration Area in Neglect: A New Analysis Method for Video-Oculography Data Based on Foveal Vision. Front Neurosci. 2019;13:1412.

21. Chiffi K, Diana L, Hartmann M, Cazzoli D, Bassetti CL, Muri RM, Eberhard-Moscicka AK. Spatial asymmetries (“pseudoneglect”) in free visual exploration-modulation of age and relationship to line bisection. Exp Brain Res. 2021;239(9):2693–700.

22. Brozzoli C, Dematte ML, Pavani F, Frassinetti F, Farne A. Neglect and extinction: within and between sensory modalities. Restor Neurol Neurosci. 2006;24(4-6):217–32.

23. Foulsham T, Underwood G. What can saliency models predict about eye movements? Spatial and sequential aspects of fixations during encoding and recognition. J Vis. 2008;8(2):6 1–17.

24. Seghier ML, Ramlackhansingh A, Crinion J, Leff AP, Price CJ. Lesion identification using unified segmentation-normalisation models and fuzzy clustering. Neuroimage. 2008;41(4):1253–66.

25. Buxbaum LJ, Ferraro MK, Veramonti T, Farne A, Whyte J, Ladavas E, et al. Hemispatial neglect: Subtypes, neuroanatomy, and disability. Neurology. 2004;62(5):749–56.

26. Bailey MJ, Riddoch MJ, Crome P. Test-retest stability of three tests for unilateral visual neglect in patients with stroke: Star Cancellation, Line Bisection, and the Baking Tray Task. Neuropsychological Rehabilitation. 2004;14(4):403–19.

27. Zeltzer L M, A. Star Cancellation Test Assessment: Section. In: Stroke Engine Assess. Montreal. 2008 [Available from: https://strokengine.ca/en/assessments/star-cancellation-test/.

28. Demeyere N, Riddoch MJ, Slavkova ED, Bickerton WL, Humphreys GW. The Oxford Cognitive Screen (OCS): validation of a stroke-specific short cognitive screening tool. Psychol Assess. 2015;27(3):883–94.

29. McDermott A. Catherine Bergego Scale (CBS) Assessment: Section. In: Stroke Engine Assess. Montreal 2012 [Available from: https://strokengine.ca/en/assessments/catherine-bergego-scale-cbs/.

30. Kaufmann BC, Cazzoli D, Pflugshaupt T, Bohlhalter S, Vanbellingen T, Muri RM, et al. Eyetracking during free visual exploration detects neglect more reliably than paper-pencil tests. Cortex. 2020;129:223–35.

31. Orland K. Hands-on: Valve/HTC Vive opens up the virtual reality experience Ars Technica 2015 [Available from: https://arstechnica.com/gaming/2015/03/hands-on-valvehtc-vive-opens-up-the-virtual-reality-experience/.

32. Halligan PW, Marshall JC. Is Neglect (Only) Lateral a Quadrant Analysis of Line Cancellation. J Clin Exp Neuropsyc. 1989;11(6):793–8.

33. Shelton PA, Bowers D, Heilman KM. Peripersonal and Vertical Neglect. Brain. 1990;113:191–205.

